# Efficacy of universal masking for source control and personal protection from simulated cough and exhaled aerosols in a room

**DOI:** 10.1101/2021.04.21.21255880

**Authors:** William G. Lindsley, Donald H. Beezhold, Jayme Coyle, Raymond C. Derk, Francoise M. Blachere, Theresa Boots, Jeffrey S. Reynolds, Walter G. McKinney, Erik Sinsel, John D. Noti

**Affiliations:** Health Effects Laboratory Division, National Institute for Occupational Safety and Health, Centers for Disease Control and Prevention, Morgantown, West Virginia, USA

**Author notes:** Corresponding author: Dr. William G. Lindsley, National Institute for Occupational Safety and Health (NIOSH), 1000 Frederick Lane, M/S 4020, Morgantown, WV 26508-5402.

**Keywords:** Infection control, Airborne transmission, Infectious disease transmission, Face masks

## Abstract

Face masks reduce the spread of infectious respiratory diseases such as COVID-19 by blocking aerosols produced during coughs and exhalations (“source control”). Masks also slow and deflect cough and exhalation airflows, which changes the dispersion of aerosols. Factors such as the directions in which people are facing (orientation) and separation distance also affect aerosol dispersion. However, it is not clear how masking, orientation, and distance interact. We placed a respiratory aerosol simulator (“source”) and a breathing simulator (“recipient”) in a 3 m x 3 m chamber and measured aerosol concentrations for different combinations of masking, orientation, and separation distance. When the simulators were front-to-front during coughing, masks reduced the 15-minute mean aerosol concentration at the recipient by 92% at 0.9 and 1.8 m separation. When the simulators were side-by-side, masks reduced the concentration by 81% at 0.9 m and 78% at 1.8 m. During breathing, masks reduced the aerosol concentration by 66% when front-to-front and 76% when side-by-side at 0.9 m. Similar results were seen at 1.8 m. When the simulators were unmasked, changing the orientations from front-to-front to side-by-side reduced the cough aerosol concentration by 59% at 0.9 m and 60% at 1.8 m. When both simulators were masked, changing the orientations did not significantly change the concentration at either distance during coughing or breathing. Increasing the distance between the simulators from 0.9 m to 1.8 m during coughing reduced the aerosol concentration by 25% when no masks were worn but had little effect when both simulators were masked. During breathing, when neither simulator was masked, increasing the separation reduced the concentration by 13%, which approached significance, while the change was not significant when both source and recipient were masked. Our results show that universal masking reduces exposure to respiratory aerosol particles regardless of the orientation and separation distance between the source and recipient.

## INTRODUCTION

People who are infected with severe acute respiratory syndrome coronavirus 2 (SARS-CoV-2), the virus that causes coronavirus disease 2019 (COVID-19), can generate aerosols of respiratory fluids containing the virus when they cough, breathe, talk, sing and sneeze.^[1-5]^ Even individuals who are asymptomatic or presymptomatic appear to be able to shed enough SARS-CoV-2 virus to infect others; one study found that a large percentage of COVID-19 infections were asymptomatic, and a second estimated that as many as half of COVID-19 infections could be the result of transmission from people with no reported symptoms.^[6, 7]^ The importance of asymptomatic and presymptomatic transmission during the COVID-19 pandemic led to recommendations by the Centers for Disease Control and Prevention (CDC) and other public health organizations for the use of face masks by everyone in public places (called universal masking), along with other measures such as physical distancing, increasing room ventilation and avoiding unnecessary indoor gatherings and crowded outdoor spaces.^[8-10]^

The primary purpose of face masks is to reduce the expulsion of droplets and aerosols containing SARS-CoV-2 into the environment (called source control).^[8, 10]^ Studies using healthy human subjects have shown that cloth face masks partially block respiratory aerosols produced during coughing, breathing and talking.^[11, 12]^ Two studies of patients with respiratory infections found that medical face masks reduced the dispersion of potentially infectious aerosols.^[13, 14]^ Although face masks do not protect the wearer from airborne particles as effectively as a respiratory protective device such as a N95 filtering facepiece respirator, they do reduce the wearer’s exposure to infectious droplets and aerosols.^[8, 15-17]^ A study in a simulated classroom setting estimated that if well-fitting masks were worn by everyone in the room, the probability of infection was reduced more than 100 times below the probability of infection when no masks were worn.^[18]^

Two quantitative source control studies by our group using an aerosol simulator found that cloth face masks and medical masks typically blocked 40%-60% of coughed and exhaled aerosol particles and were more effective as the particle size increased.^[19, 20]^ In a subsequent study, we found that if a medical mask was knotted and the pleats were tucked to improve the fit of the mask, the mask blocked 77% of the aerosol particles, and the aerosol blocking could be increased to 85% by wearing a cloth mask over a medical mask (double masking).^[21]^ We examined the efficacy of universal masking by placed a respiratory aerosol simulator (simulating a “source” person coughing or exhaling respiratory aerosols) and a breathing simulator (simulating a “recipient” person inhaling the aerosols) in a small room with aerosol particle measurement instruments.^[21]^ When both the source and recipient simulators were wearing a well-fitting medical mask or a cloth mask over a medical mask, the exposure of the recipient to simulated respiratory aerosols was reduced by 96%.

Several epidemiological studies conducted during the current pandemic have concluded that universal masking helps reduce COVID-19 transmission.^[8]^ A comparison of counties with and without mask mandates in the US state of Kansas found that mask mandates were associated with lower incidence rates of COVID-19.^[22]^ Similar outcomes were seen after implementation of face mask mandates in Germany.^[23]^ Implementation of multiple infection reduction measures, including universal masking, led to a significantly lower rate of SARS-CoV-2 among healthcare workers in a hospital system.^[24]^ A study of fifteen US states and the city of Washington, DC, found that face mask mandates were associated with a decline in the growth rate of COVID-19 cases,^[25]^ while a study of ten states found that statewide mask mandates were associated with a reduction in the growth rate of COVID-19 hospitalizations.^[26]^

The purpose of this study was to characterize the influence of various factors on the efficacy of universal face mask use. We used a respiratory aerosol simulator (source) to cough or exhale aerosols into a chamber and measured the aerosol concentration over time in the breathing zone of a breathing simulator (recipient). Experiments were conducted with no masks or cloth masks on the source and recipient to determine how different combinations of masking, simulator orientations, and separation distances affected the aerosol exposure of the recipient. Our results help provide information to the public health community to better understand the potential effect of universal masking on the transmission of respiratory infections like SARS-CoV-2.

## METHODS

### Respiratory aerosol source simulator

A respiratory aerosol source simulator was used to simulate a person (called the source) who was coughing or exhaling aerosol particles into the test chamber. The source simulator was based on a system used to test masks as source control devices for respiratory aerosols.^[19, 20]^ The simulator uses an elastomeric bellows driven by a computer-controlled linear motor to reproduce human coughing and breathing airflows. The source simulator includes a manikin headform that has pliable skin mimicking the elastic properties of human skin in order to create a realistic simulation of how each source control device would fit a human face.^[27]^ The mouth of the respiratory aerosol simulator was 1.5 m (60”) above the floor of the chamber.

The test aerosol was produced using a solution of 14% potassium chloride (KCl) in a single-jet Collison nebulizer (BGI, Butler, NJ) at 103 kPa (15 lbs./in^2^). The aerosol passed through a diffusion drier (Model 3062, TSI, Shoreview, MN), mixed with dry filtered air flowing at 10 L/min for the cough tests and 15 L/min for the breathing tests, and was neutralized using a bipolar ionizer (Model HPX-1, Electrostatics). For cough aerosol tests, the test aerosol was loaded into the elastomeric bellows and then expelled using a single cough with a volume of 4.2 L and a peak flow rate of 11 L/s.^[28]^ For breathing tests, the system used a ventilation rate of 15 L/min with a breathing rate of 12 breathes/min and a tidal volume of 1.25 liters, which corresponds to the ISO standard for a female performing light work.^[29]^ During the breathing experiments, the nebulizer was continuously cycled on for 10 seconds and off for 50 seconds to prevent the aerosol concentration in the chamber from exceeding the upper concentration limit of the aerosol particle counters.

### Breathing recipient simulator

A digital breathing simulator (Warwick Technologies Ltd., Warwick, UK) with a pliable skin headform (Respirator Testing Head Form 1 – Static, Crawley Creatures Ltd, Buckingham, UK) was used to simulate a person (called the recipient) who was in the room and exposed to the respiratory aerosol particles expelled by the source simulator. The breathing waveform was sinusoidal with a breathing rate of 21.5 breaths/minute and a ventilation rate of 27 liters/minute, which is approximately the average of the ISO standards for males and females engaged in moderate work.^[29]^ The mouth of the breathing simulator was 1.5 m above the floor of the chamber.

### Face masks and fit testing

The face mask used in our experiments was a cloth face mask with 3 layers of cotton fabric and ear loops (Defender model, HanesBrands Inc., Winston Salem, NC). Experiments were conducted with four masking conditions: (1) No masks on either the source or receiver (No mask/no mask); (2) A mask on the receiver only (No mask/cloth mask); (3) A mask on the source only (Cloth mask/no mask); and (4) Masks on both the source and receiver (Cloth mask/cloth mask). After the mask was placed on the headform, the fit factor was measured as described previously^[20]^ using a standard respirator fit testing device (Model 8038 PortaCount® Pro Plus; TSI). The PortaCount was used with the N95 companion method, which counts negatively-charged particles 55 nm in diameter.^[30, 31]^ The fit factor (FF) was calculated as:^[32, 33]^

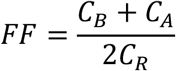

Where

C_B_ = particle concentration in the ambient sample taken before the mask sample.

C_A_ = particle concentration in the ambient sample taken after the mask sample.

C_R_ = particle concentration in the sample from inside the mask.

### Aerosol particle concentration

During experiments, aerosol particle concentrations at six locations in the exposure chamber were continuously monitored using optical particle counters (OPC, Model 1.108; Grimm Technologies, Inc., Douglasville, GA). One particle counter was located at the mouth of the breathing simulator so that, if the simulator was wearing a face mask, the particle counter collected aerosol samples from inside the mask (i.e., the particle counter measured the concentration of the aerosol being inhaled by the recipient simulator). The optical particle counters reported the number of aerosol particles detected per liter of air (#/L) at 1 Hz in eight logarithmically spaced size bins from 0.3 to 3 µm, except for one older unit which reported data at 1/6 Hz. The inlets of the aerosol particle counters were located 1.5 m above the floor of the chamber.

### Test procedure

Our experiments were conducted in a 3.2 m x 3.2 m x 2.3 m high (124” x 124” x 89”) environmental chamber with a volume of 22.5 m^3^. The chamber includes a HEPA filtration system with a 4.5 m^3^/minute flowrate. The mean chamber temperature was 74.8 °C (SD 1.5 °C), and the mean relative humidity was 26.7% (SD 2.5%) during the experiments. Experiments were performed with the source and recipient simulator either 0.9 m (36”) or 1.8 m (72”) apart, measured from the mouth opening of each simulator (Figure 1). The simulators were oriented so that they were either: (1) facing each other (front-to-front), (2) with the front of the source simulator facing the back of the recipient simulator (front-to-back), or (3) with the simulators beside each other facing in the same direction (side-by-side). Detailed schematics of the different simulator arrangements and the OPC locations are provided in the supplemental materials.

**Figure 1:**
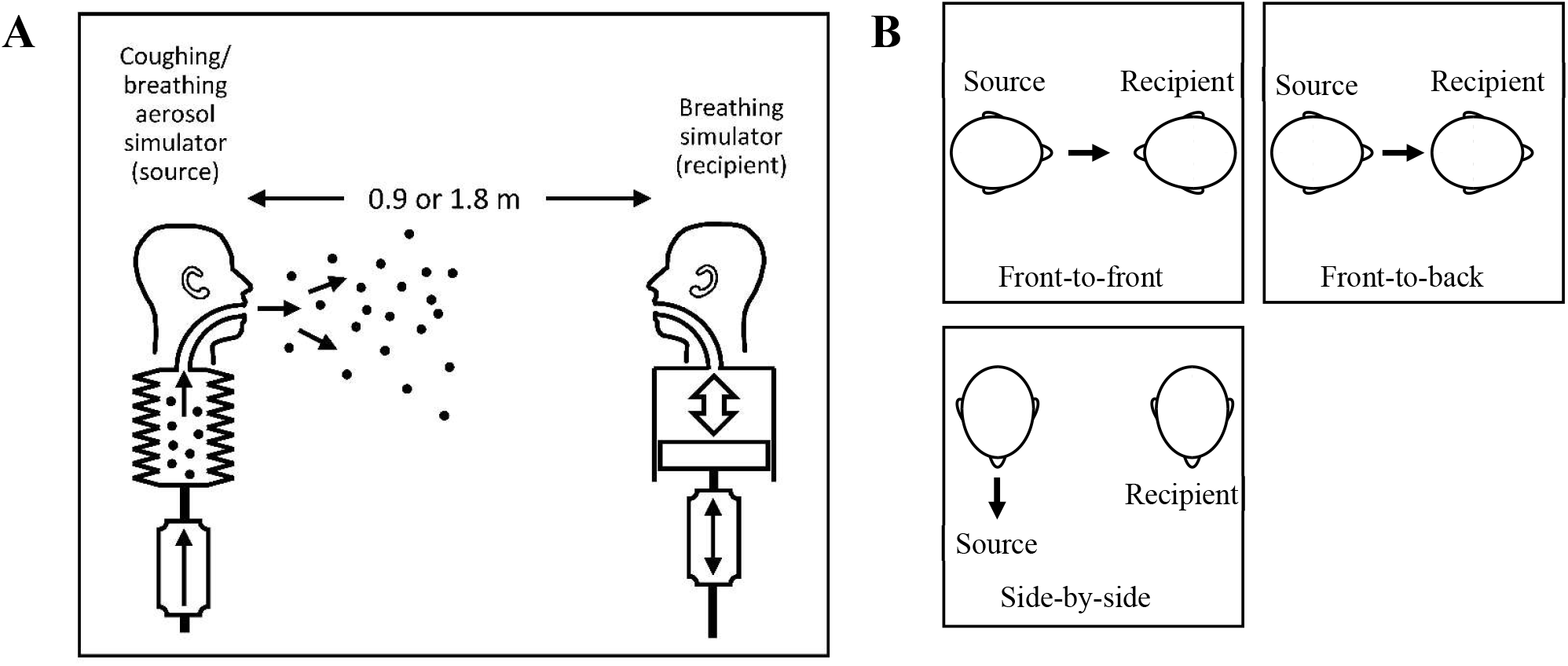
Schematic of source and recipient simulators in environmental chamber (not to scale). (A) Side view. (B) Top view showing three simulator orientations. More detailed information is shown in Figures S1 and S2 in the supplemental information.

Before each experiment, the source and recipient simulators were positioned in the chamber. Masks were placed on the source and recipient simulator as appropriate and the fit factors were measured. The chamber was sealed, and the HEPA filtration system was turned on for 30 minutes to remove airborne particles. The HEPA system was turned off 15 minutes before each experiment to reduce the air currents within the chamber, during which time the OPCs measured the background aerosol concentration. The source simulator then either coughed once into the chamber or breathed continuously. Aerosol concentrations were collected at all locations for 20 minutes. Each combination of masking, simulator orientation, simulator separation distance, and coughing or exhaling aerosol was tested three times for a total of 144 experiments. The experimental parameters are shown in Table 1.

**Table 1:** Experimental parameters

| *Experimental parameter* | *Values tested* |
| --- | --- |
| Mask on source simulator | No mask<br>Cloth mask |
| Mask on recipient simulator | No mask<br>Cloth mask |
| Respiratory activity by source simulator | Single 4.2-liter cough<br>Continuous 15 liters/minute breathing |
| Respiratory activity by recipient simulator | Continuous 27 liters/minute breathing |
| Simulator orientations | Front-to-front<br>Front-to-back<br>Side-by-side |
| Mouth-to-mouth distance | 0.9 m (36")<br>1.8 m (72") |

### Data Analysis

The background aerosol number concentration was calculated based on the mean number concentration in each size bin during the three minutes before the cough or exhalation by the source simulator and was subtracted from the concentrations measured afterward. The mass of the aerosol in each size bin per m^3^ of air (mass concentration) was calculated by multiplying the particle count by the volume of an individual particle with the mean diameter of the size bin (assuming the particles were spherical) and by 1.984 g/cm^3^ (the density of KCl). Note that this conversion from particle counts to particle mass is commonly used but is an approximation. The total aerosol mass/m^3^ (total aerosol mass concentration) was found by summing the aerosol mass concentrations for all the size bins. The mean mass concentration was found by averaging the total mass concentration over 15 minutes starting from the time of the cough or initial exhalation of the test aerosol.

The data were analyzed via an ANOVA for within-Orientation, within-Condition, and between-Distance comparisons (blocking for orientation in between-Distance). All analyses were completed in R with ‘lsmeans’ and ‘lmerTest’ packages.^[34-36]^

## RESULTS

An example of the aerosol particle concentration measured at the mouth of the recipient simulator during a coughing experiment is shown in Figure 2. Because the entire cough aerosol is expelled in 1.2 seconds, the concentration can be seen to rise rapidly at the beginning of the collection period and then fall off as the aerosol disperses in the room. This effect was most pronounced when the source and recipient simulator were facing each other. Placing a mask on the source simulator greatly attenuated the initial spike in concentration. An example of the particle concentration during a breathing experiment is shown in Figure 3. In this case, because the aerosol is being exhaled over time and because the flow velocity at the mouth of the source simulator is much lower, the aerosol concentration at the mouth of the recipient increases steadily.

**Figure 2:**
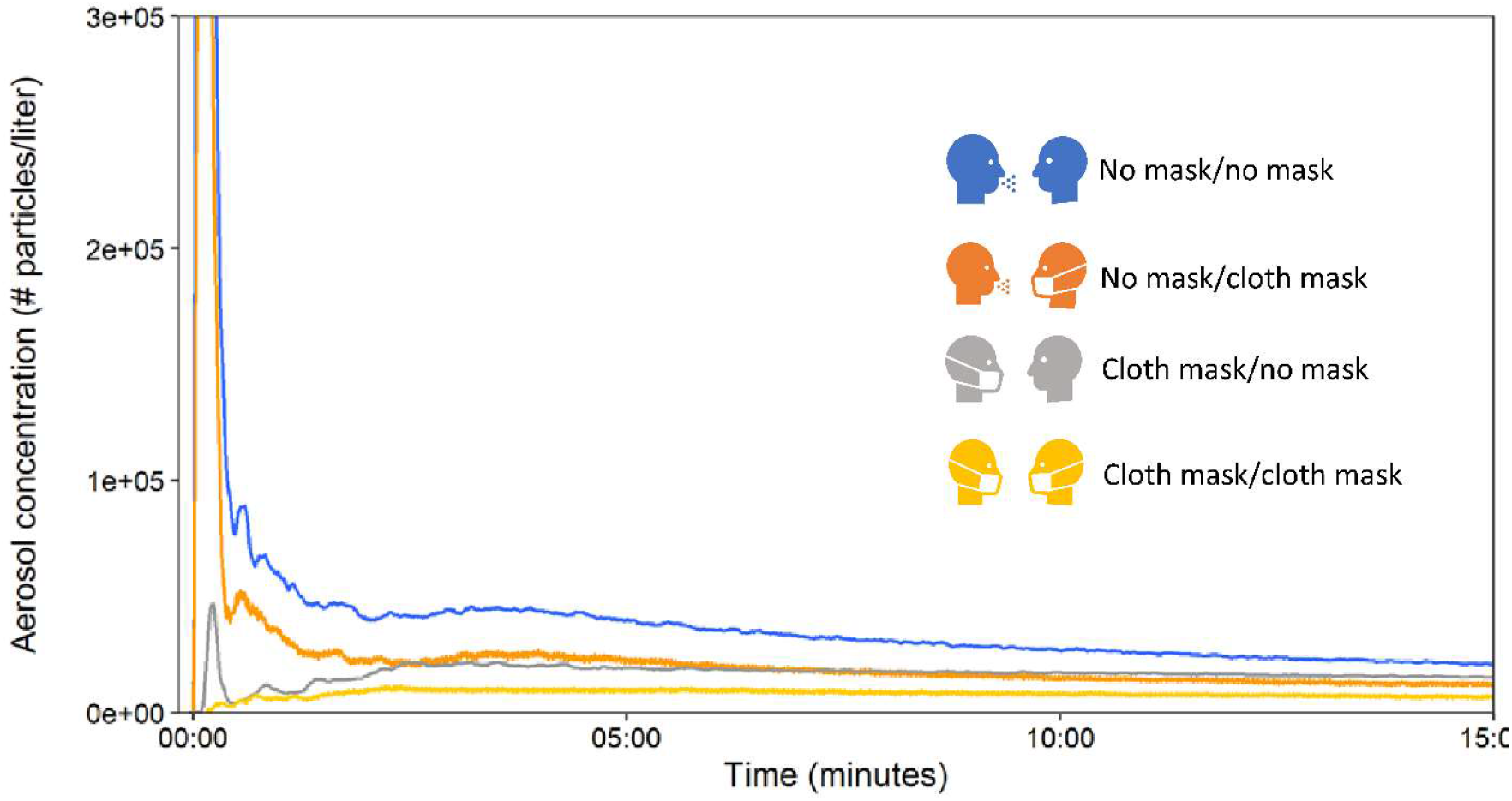
Aerosol concentration at mouth of recipient (breathing simulator) after single cough from source (cough & exhaled aerosol simulator). The concentration is shown as total particles/liter for 0.3 to 3 µm aerosol particles, which is the format reported by the aerosol particle counter. Source and recipient were 1.8 m (72”) apart and facing each other. Each line shows data from a single experiment. The plots were smoothed with a 7-point running average for clarity.

**Figure 3:**
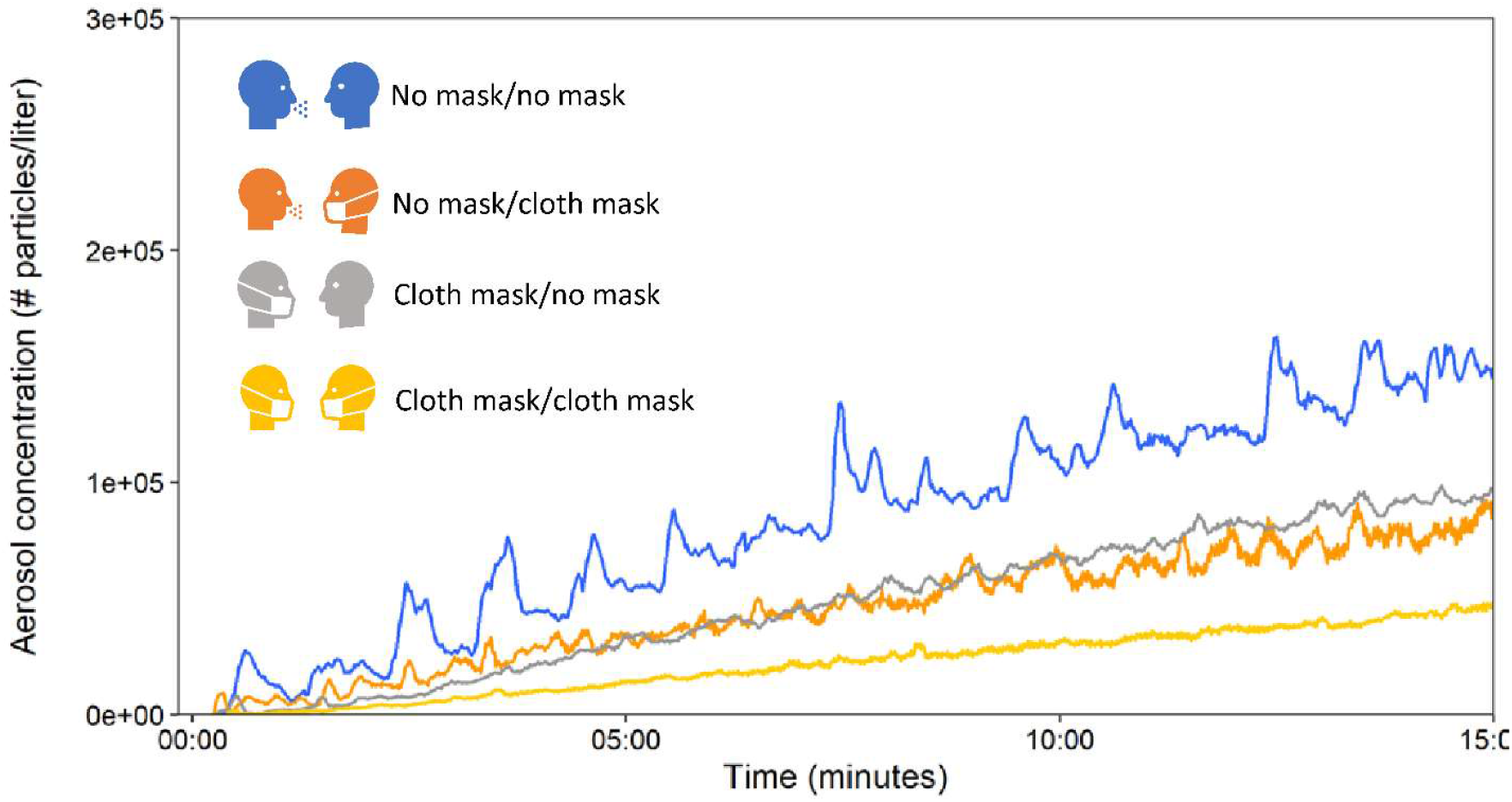
Aerosol concentration at mouth of recipient (breathing simulator) while source (cough & exhaled aerosol simulator) is exhaling aerosol. Source and recipient were 1.8 m (72”) apart and facing each other. Each line shows data from single experiment. The plots were smoothed with a 7-point running average for clarity.

The mean aerosol concentrations at the mouth of the recipient simulator during coughing and breathing experiments with the simulators 0.9 m and 1.8 m apart are shown in Table 2 and Figure 4. The experiments were conducted with the simulators facing each other (front-to-front), with the source simulator facing the back of the recipient simulator (front-to-back), and with the simulators facing the same direction (side-by-side). The average fit factor for masks was 1.6 (SD 0.4) on the source simulator and 4.6 (SD 2.5) on the recipient simulator. The use of face masks led to a reduction in concentration during both coughing and breathing experiments at both separation distances when compared with the corresponding no mask tests. This effect was most pronounced when the two simulators were front-to-front during the coughing experiments; at both distances, a mask on the source reduced the concentration by 80% (p < 0.0001), while a mask on the recipient reduced the concentration by 41% at a 0.9 m separation (p = 0.0001) and 48% at 1.8 m (p = 0.0002), and masks on both source and recipient reduced the concentration by 92% (p < 0.0001) at both 0.9 and 1.8 m. Similar results were seen for the other orientations, although the effects of masking were not as strong. For the side-by-side coughing experiments, placing a mask on the source reduced exposure by 46% at 0.9 m (p < 0.0001) and 38% at 1.8 m (p = 0.0002), while placing masks on both the source and recipient reduced the concentration by 81% at 0.9 m and 78% at 1.8 m (p < 0.0001 for both). For the breathing experiments, placing masks on both simulators at a 0.9 m separation reduced the aerosol concentration by 66% when front-to-front (p = 0.0061), 78% when front-to-back (p < 0.0001), and 76% when side-by-side (p < 0.0001). Similar results were seen at a 1.8 m separation.

**Table 2:**
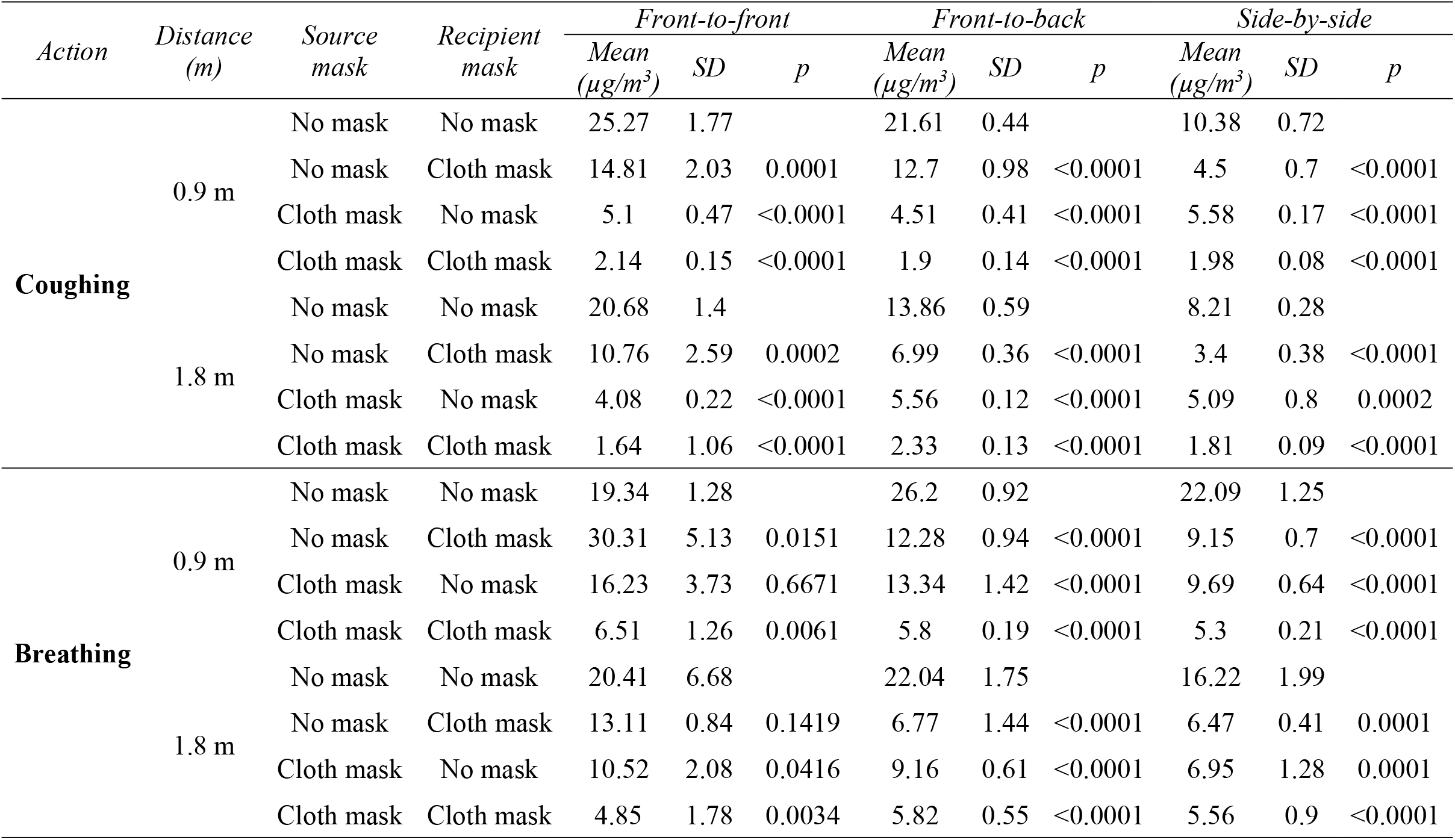
Mean aerosol concentrations over 15 minutes for coughing and breathing experiments. The p-values are for comparisons of the experiments with no masks on the source and recipient to the other masking conditions with each block of action, orientation, and distance evaluated separately.

**Figure 4:**
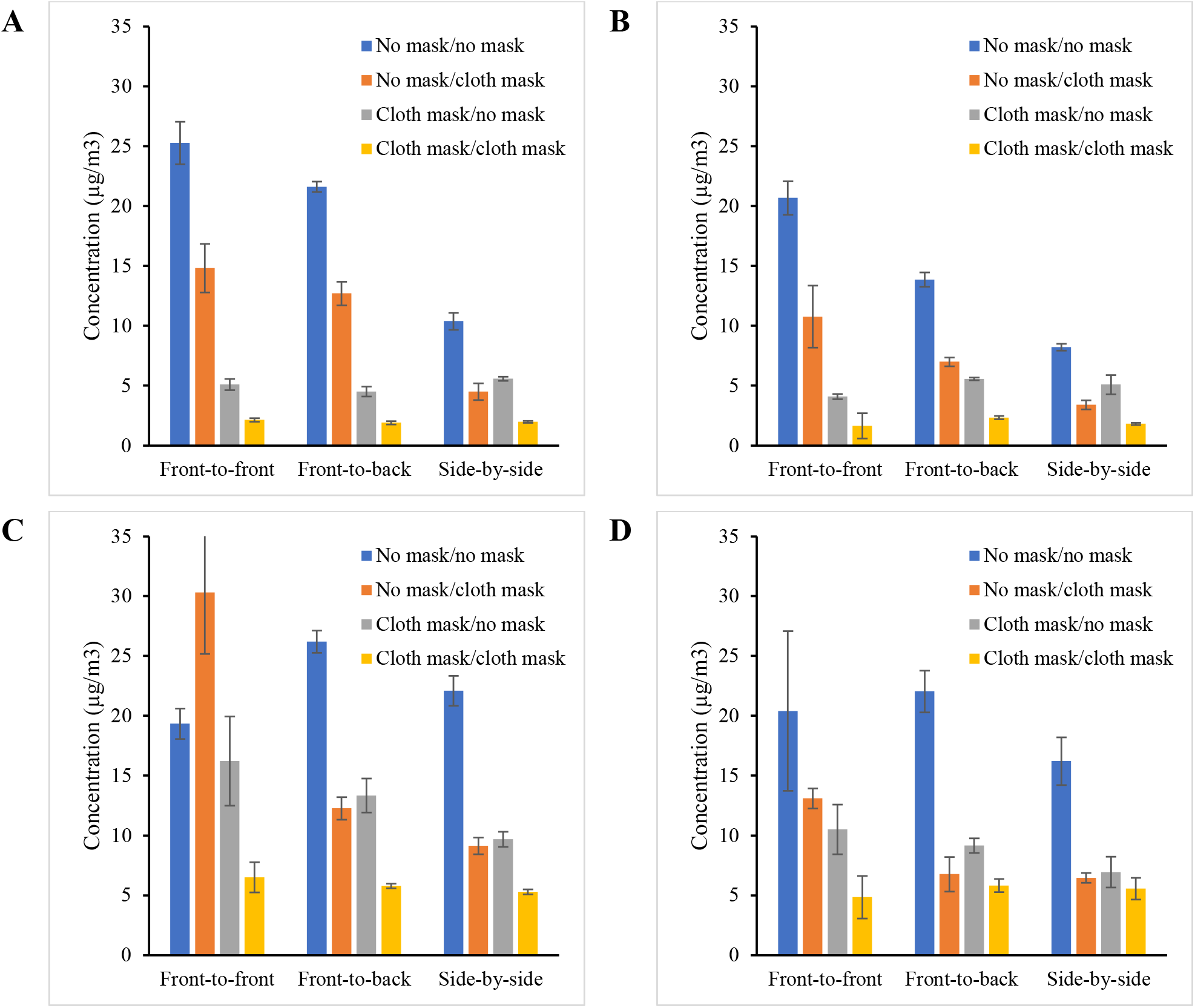
Mean aerosol concentration over 15 minutes measured at the mouth of the recipient simulator with and without masks and with simulators in different orientations. (A) Source is coughing, simulators are 0.9 m (36”) apart. (B) Source is coughing, simulators are 1.8 m (72”) apart. (C) Source is breathing, simulators are 0.9 m apart. (D) Source is breathing, simulators are 1.8 m apart. Each bar is the mean of three experiments. Error bars show the standard deviation.

The relative orientation of the source and recipient simulators had less of an effect on the concentration at the recipient than did masking, especially when a mask was placed on the source. For the coughing experiments with no masks on the source or recipient, changing the simulator orientations from the front-to-front orientation to front-to-back reduced the concentration by 15% (p = 0.0178; Table 3) with a 0.9 m separation and 33% (p = 0.0001) with a 1.8 m separation. When the orientation was changed from the front-to-front orientation to side-by-side, the concentration decreased by 59% at 0.9 m and 60% at 1.8 m (p < 0.0001 for both). On the other hand, for cough experiments in which masks were worn by both the source and recipient, changing the orientations from front-to-front to either front-to-back or side-by-side did not significantly change the concentration at either separation distance (p = 0.1176 to 0.9428). For the breathing experiments in which no masks were worn at a separation of 0.9 m, changing the orientation from front-to-front to front-to-back increased the concentration by 36% (p = 0.0009) and changing from front-to-front to side-to-side increased it by 14% (p = 0.0624, approached significance). Neither change in orientation resulted in a significant change in concentration at 1.8 m (p = 0.8815 and 0.4762). When masks were worn by both the source and recipient, changing the simulator orientation did not significantly change the concentration at either distance (p = 0.1958 to 0.7570).

**Table 3:** P-values for comparisons of different simulator orientations. Each block of action, distance, and masking conditions was evaluated separately.

| *Action* | *Distance*<br>*(m)* | *Source*<br>*mask* | *Recipient*<br>*mask* | *Front-to-front vs.*<br>*front-to-back*<br>*(p)* | *Front-to-front vs.*<br>*side-by-side*<br>*(p)* |
| --- | --- | --- | --- | --- | --- |
| **Coughing** | 0.9 m | No mask | No mask | 0.0178 | <0.0001 |
|  |  | No mask | Cloth mask | 0.2188 | 0.0002 |
|  |  | Cloth mask | No mask | 0.2039 | 0.3278 |
|  |  | Cloth mask | Cloth mask | 0.1176 | 0.3085 |
|  | 1.8 m | No mask | No mask | 0.0001 | <0.0001 |
|  |  | No mask | Cloth mask | 0.0530 | 0.0026 |
|  |  | Cloth mask | No mask | 0.0219 | 0.0946 |
|  |  | Cloth mask | Cloth mask | 0.4147 | 0.9428 |
| **Breathing** | 0.9 m | No mask | No mask | 0.0009 | 0.0624 |
|  |  | No mask | Cloth mask | 0.0008 | 0.0003 |
|  |  | Cloth mask | No mask | 0.3494 | 0.0321 |
|  |  | Cloth mask | Cloth mask | 0.5174 | 0.1958 |
|  | 1.8 m | No mask | No mask | 0.8815 | 0.4762 |
|  |  | No mask | Cloth mask | 0.0006 | 0.0004 |
|  |  | Cloth mask | No mask | 0.5247 | 0.0537 |
|  |  | Cloth mask | Cloth mask | 0.6066 | 0.7570 |

For all orientations combined, increasing the distance between the simulators from 0.9 m to 1.8 m during the coughing experiments reduced the aerosol concentration by 25% when no masks were used (Table 4; p < 0.0001) and 34% when the recipient was masked (p = 0.0005). When the source was masked, distance had less effect on the concentration when the recipient was unmasked (3% decrease, p = 0.6157) or when both were masked (4% decrease, p = 0.7084). For the breathing experiments, when the source was unmasked and the recipient was masked, increasing the separation reduced the concentration by 49% (p = 0.0007). When both simulators were unmasked, the concentration decreased by 13%, which approached significance (p = 0.0737). When the source was masked and the recipient unmasked, the decrease was 32% (p = 0.0004), while the change was not significant when both source and recipient were masked (8% decrease, p = 0.3403).

**Table 4:** Mean aerosol concentrations at recipient at 0.9 m and 1.8 m. Each value is the mean of all orientations for each distance and masking condition. The p-values compare the results at 0.9 m with the results at 1.8 m for each masking condition.

| *Action* | *Source mask* | *Recipient mask* | *0.9 m distance* |  | *1.8 m distance* |  | *p* |
| --- | --- | --- | --- | --- | --- | --- | --- |
|  |  |  | *Mean ($\mu\text{g}/\text{m}^3$)* | *SD* | *Mean ($\mu\text{g}/\text{m}^3$)* | *SD* |  |
| **Coughing** | No mask | No mask | 19.09 | 6.79 | 14.39 | 5.65 | <0.0001 |
|  | No mask | Cloth mask | 10.67 | 4.86 | 7.05 | 3.45 | 0.0005 |
|  | Cloth mask | No mask | 5.07 | 0.57 | 4.91 | 0.78 | 0.6157 |
|  | Cloth mask | Cloth mask | 2.01 | 0.15 | 1.92 | 0.62 | 0.7084 |
| **Breathing** | No mask | No mask | 22.54 | 3.15 | 19.56 | 4.44 | 0.0737 |
|  | No mask | Cloth mask | 17.25 | 10.23 | 8.78 | 3.36 | 0.0007 |
|  | Cloth mask | No mask | 13.09 | 3.48 | 8.88 | 2.01 | 0.0004 |
|  | Cloth mask | Cloth mask | 5.87 | 0.83 | 5.41 | 1.12 | 0.3403 |

In addition to measuring the aerosol concentration at the mouth of the recipient, aerosol concentration data were collected at five other locations in the chamber during our experiments. The mean concentrations varied from location to location depending upon masking, the orientation and location of the source and recipient, and whether the source was coughing or breathing. The mean aerosol concentrations for the front-to-front and side-by-side orientations when the simulators were 1.8 m apart are shown in Figure 5. To examine the effects of placing a face mask on the source simulator during coughing and breathing, the measurements from the five locations other than the mouth of the recipient simulator were averaged to calculate an aerosol concentration for the entire chamber when the source simulator was unmasked or masked (Figure 6). Placing a mask on the source reduced the chamber concentration by a range of 29% to 69% during cough experiments and 30% to 61% during breathing experiments, and the differences were significant in all cases (Table 5).

**Table 5:** Overall chamber mean aerosol concentrations calculated using data from five locations. at recipient at 0.9 m and 1.8 m. The p-values compare the results with no mask on the source to the results with the source wearing a cloth mask. The recipient was unmasked for all experiments.

| Action | Distance | Orientation | Source masking |  |  |  | p |
| --- | --- | --- | --- | --- | --- | --- | --- |
|  |  |  | No mask |  | Cloth mask |  |  |
| | | | Mean<br>($\mu\text{g}/\text{m}^3$) | SD | Mean<br>($\mu\text{g}/\text{m}^3$) | SD | |
| Coughing | 0.9 | Front-to-front | 16.00 | 1.58 | 6.07 | 0.09 | <0.0001 |
|  |  | Front-to-back | 18.84 | 0.45 | 5.80 | 0.56 | <0.0001 |
|  |  | Side-by-side | 11.30 | 0.70 | 6.47 | 0.77 | 0.0001 |
|  | 1.8 | Front-to-front | 15.60 | 0.74 | 6.02 | 0.08 | <0.0001 |
|  |  | Front-to-back | 14.40 | 0.48 | 7.67 | 0.58 | 0.0001 |
|  |  | Side-by-side | 10.22 | 0.42 | 7.30 | 1.38 | 0.0149 |
| Breathing | 0.9 | Front-to-front | 31.65 | 2.44 | 22.04 | 4.55 | 0.0376 |
|  |  | Front-to-back | 39.66 | 2.37 | 22.89 | 3.88 | 0.0001 |
|  |  | Side-by-side | 24.34 | 1.37 | 11.69 | 1.07 | <0.0001 |
|  | 1.8 | Front-to-front | 36.85 | 5.83 | 17.66 | 3.03 | 0.0026 |
|  |  | Front-to-back | 46.13 | 0.71 | 20.97 | 2.10 | <0.0001 |
|  |  | Side-by-side | 26.47 | 2.72 | 10.21 | 1.28 | 0.0014 |

**Figure 5:**
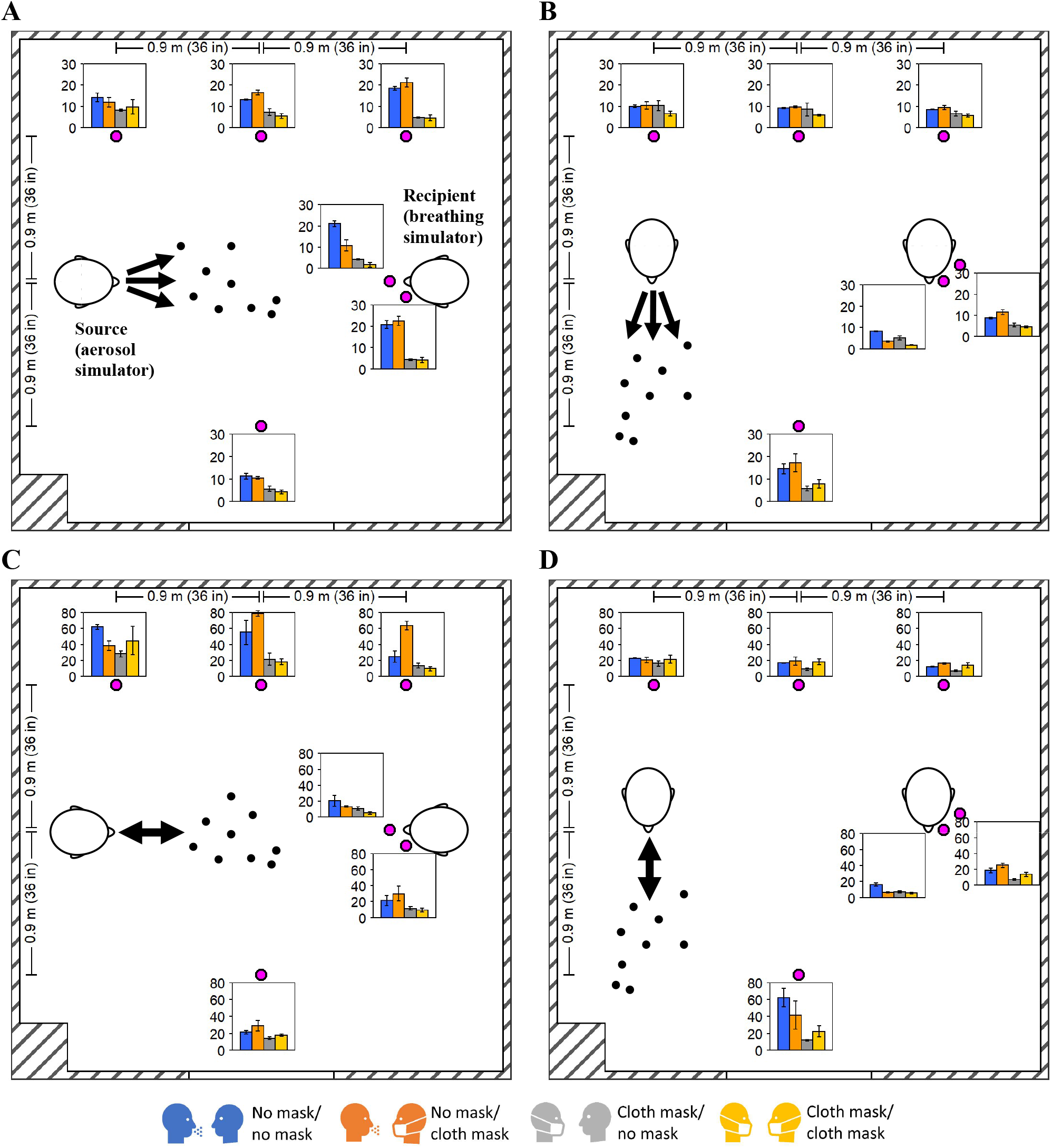
Mean aerosol concentrations at different locations in chamber with simulators 1.8 m apart. The magenta dots indicate the particle counter locations. The bar plots show the mean concentrations at each location with masking as indicated in the legend below the plot. The mean values are reported in µg/m^3^ and were calculated over 15 minutes. Error bars indicate the standard deviations for three experiments. (A) Source is coughing, simulators are front-to-front. (B) Source coughing, simulators side-by-side. (C) Source breathing, simulators front-to-front. (D) Source breathing, simulators side-by-side. Additional results are shown in Figures S3 and S4 in the supplemental information.

**Figure 6:**
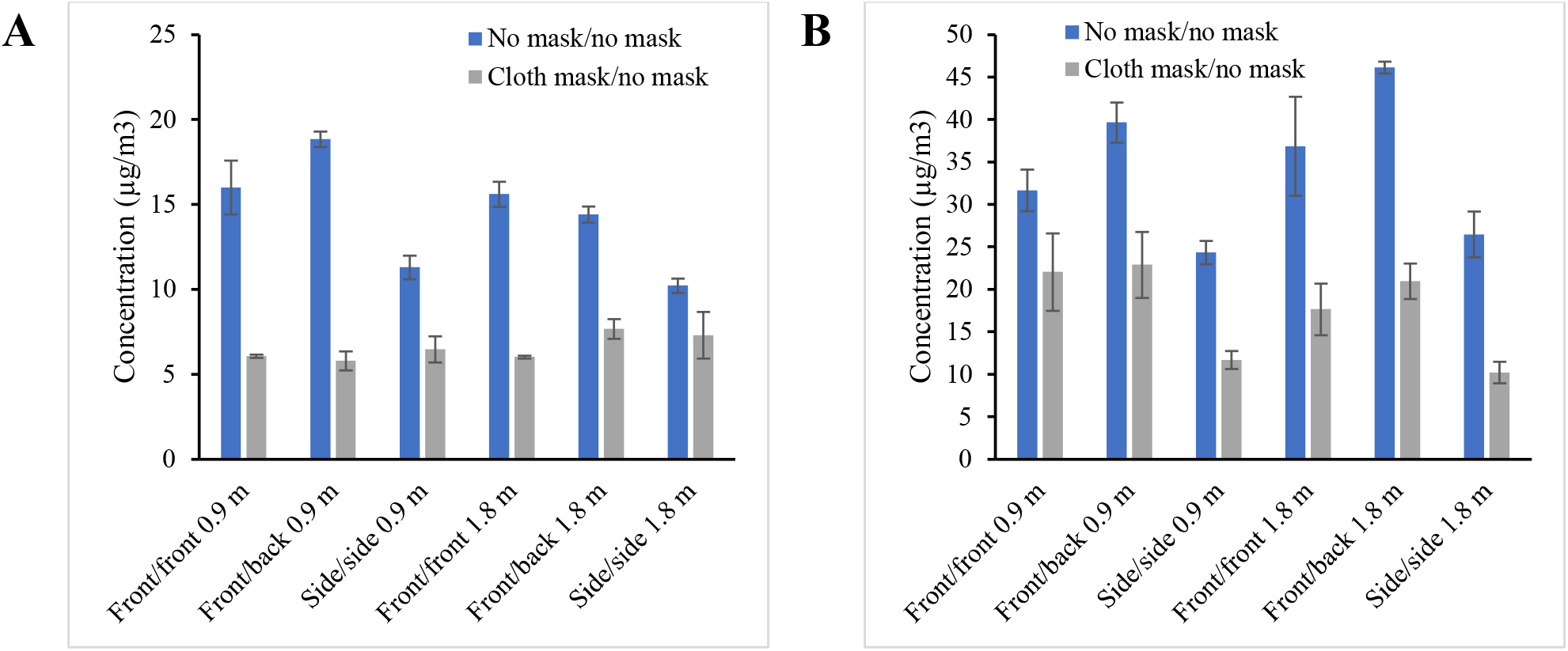
Aerosol concentration in the environmental chamber with and without a mask on the source simulator during (A) coughing experiments and (B) breathing experiments. The recipient simulator was unmasked during these experiments. The simulators were 0.9 or 1.8 m apart and oriented front-to-front, front-to-back, or side-by-side. For each experiment, the 15-minute mean concentrations at five locations (excluding the location at the mouth of the recipient) were averaged to determine the chamber concentration. Each bar shows the average and standard deviation of the chamber concentrations from three experiments. Note that the source simulator expels a larger mass of aerosol during breathing experiments than during coughing experiments.

## DISCUSSION

The use of face masks by everyone in public spaces is recommended to help reduce the transmission of SARS-CoV-2 by reducing the exposure of uninfected individuals to infectious aerosols and droplets.^[8-10]^ The most important benefit of mask wearing is blocking aerosol particles from coughs and exhalations, which reduces the amount of aerosol that is released into the environment. Previously, we have shown that the cloth mask used here reduced the expulsion of cough aerosols by 52% and exhalation aerosols by 44%.^[20]^ However, universal masking can also have less obvious effects that can affect the exposure of others to respiratory aerosols. The mask on the source slows and disperses the airstream passing through the mask, which reduces the projection of a cough or exhalation jet into a room, and the mask can deflect part of a cough or exhalation.^[37]^ Thus, in addition to reducing the aerosol concentration by filtration, placing a mask on the source changes the rate of dispersion and the spatial distribution of the respiratory aerosols in a room. Similarly, placing a mask on the recipient removes some of the particles from the inhaled air depending upon the filtration efficiency of the mask and how well it fits the recipient.^[15, 16]^ In addition, though, the recipient’s mask affects the inhalation and exhalation airflow patterns around the recipient, which in turn affects the flow of air (and thus aerosol concentration) around the recipient.^[37]^ In addition to the effects of mask usage, the exposure of a recipient person to respiratory aerosols can be influenced by factors such as the relative location of the source and recipient, the directions in which the source and recipient are facing, and the length of time that the source and recipient are in a room together. Together, these factors can interact in complex ways that may be difficult to predict.

Consider, for example, Figure 2, which shows the aerosol concentrations detected at the mouth of the recipient after a cough when the source and recipient were 1.8 m apart and facing each other. When the source was unmasked, a sharp initial spike in aerosol concentration occurred immediately after the cough. When the source was wearing a mask, the initial spike was largely attenuated. This attenuation occurred in part because the mask filters out much of the expelled aerosol. However, it also occurred because an unmasked cough forms a narrow jet that can carry aerosol particles in the direction of the cough, and this jet is greatly diminished by placing a mask on the source. This result can also be seen in Figure 4. When the source and recipient were front-to-front and the source was coughing, placing a mask on the source reduced the aerosol concentration at the mouth of the recipient by 80%, which is considerably more than the 52% predicted by source control performance alone. When the source and recipient were side-by-side so that the source was not coughing directly at the recipient, the concentration was reduced by only 46% at 0.9 m and 38% at 1.8 m, which is comparable to the reduction expected due to source control alone. Thus, comparing the front-to-front and side-by-side results shows the additional effect of attenuating the cough jet when the jet is impinging directly on the recipient.

The situation is somewhat different when looking at breathing rather than coughing (Figure 3). A cough forms a much stronger jet than does an exhalation; the average flowrate during a simulated cough was 210 liters/minute, while the flowrate during breathing averaged 15 liters/minute. Consequently, when the source and recipient were front-to-front, placing a mask on the source during breathing experiments only reduced the aerosol concentration by 16% at 0.9 m and by 49% at 1.8 m. The 16% reduction at 0.9 m in particular is much less than would be expected based simply on source control and suggests that the mask on the source slowed and spread the exhaled aerosol cloud such that it remained in the vicinity of the recipient rather than being carried past it and dispersing in the room. Thus, when the source and recipient were facing each other, placing a mask on the source reduced exposure more for a recipient who was further away than for a recipient who was closer. This effect was not seen at other orientations; when the simulators were side-by-side, placing a mask on the source reduced the concentration by 56% at 0.9 m and by 57% at 1.8 m, and orienting them front-to-back reduced the concentration by 49% at 0.9 m and by 58% at 1.8 m. These results suggest that placing a mask on the source alters the airflow patterns and aerosol mixing in the chamber caused by the coughing and breathing of the source and the breathing of the recipient, and that these alterations can change the transport and distribution of aerosols from the source in complex and unexpected ways. Placing a mask on the recipient also alters the airflow patterns and can lead to unexpected results; Figure 4C and Figure S3C show that masking the recipient but not the source resulted in higher aerosol concentrations around the recipient, suggesting that the dampening of the airflow from the recipient’s breathing reduced the dispersion of the aerosol from the source and led to an increase in localized aerosol levels.

Increasing the distance between the simulators decreased the concentration at the mouth of the recipient when the results for all orientations were averaged for each masking condition, although the effect was greatly reduced when the source and recipient were masked (Table 4). The reductions were significant in two of four masking combinations each for coughing and breathing, with one additional combination approaching significance for coughing. These results are especially interesting because our study was limited to small aerosol particles (0.3 to 3 µm), which settle slowly and are more easily dispersed than larger particles. For example, a particle in the middle of this size range (1.6 µm) would take 3.3 hours to fall 1 meter, while a 16 µm would fall 1 meter in 2.2 minutes. Thus, the reduction in exposure to respiratory aerosols that was seen with increasing distance would be expected to be much greater for larger particles. The SARS-CoV-2 virus and influenza virus are about 100 nm in diameter, but contagious humans do not shed bare viral particles. Instead, they expel aerosols and droplets of respiratory fluids that contain respiratory viruses, and the size of these virus-laden aerosols and droplets can range from hundreds of nanometers to visible droplets of 1 mm or more.^[38, 39]^ A previous study by our group using a cough aerosol with a volume median diameter of 8.5 µm showed that increasing the separation distance from 0.46 m to 1.8 m decreased exposure by 92%,^[40]^ and an experimental and modeling study found that while larger droplets tend to quickly settle out of the turbulent cloud produced by coughs and sneezes, smaller aerosols remain entrained in the cloud and can be carried several meters from the source.^[41]^

The distribution of aerosols within the chamber during coughing and breathing experiments at 1.8 m is shown in Figure 5. In all cases, the respiratory aerosol was dispersed throughout the chamber over the course of the experiment, and a person anywhere in the chamber would have been exposed to aerosol particles. The concentrations at the different locations are generally lower with masking, but in some locations, placing a mask on the recipient but not the source led to no change or a small increase rather than decrease in concentration. This presumably occurred because the changes in the velocity field around the recipient’s mouth reduced the aerosol mixing and altered the transport paths for the aerosol throughout the chamber. Although this result would not necessarily be expected, it is not unreasonable given the aerosol concentration gradients within the chamber and the changes in the air velocity fields caused by masking, and it again demonstrates that the dispersion of the respiratory aerosol within the chamber is more complex than may be appreciated from a casual analysis. However, despite some exceptions at individual locations, the overall average of the aerosol concentrations in the chamber was consistently reduced by placing masks on the source and recipient regardless of simulator orientation, separation distance, or whether the source was coughing or breathing (Figure 6 and Table 5).

Our study has several limitations. First, the optical particle counters measured airborne particles from 0.3 to 3 µm, which is a size range that includes bioaerosol particles that are small enough to remain airborne for an extended time but large enough to carry pathogens. However, humans produce aerosol particles across a broad size distribution,^[38, 39]^ and particles outside the size range in our experiments would behave differently. Second, the source and recipient simulators and the cough and exhaled aerosols were at room temperature, not body temperature. Air currents created by plumes of warm air from cough and exhalations or rising from the body can lift aerosol particles and extend the time for which they stay in the air, which could increase exposure to respiratory aerosols.^[42]^ Third, the upper detection limit of the aerosol particle counters is 2 x 10^6^ particles/liter. This limit was exceeded at the mouth of the recipient and next to the recipient headform during the first several seconds of the coughing experiments when the source was unmasked (that is, during the spike in concentration seen in Figure 2) and thus those concentration measurements were likely too low during that time. Fourth, the particle counters were not uniformly distributed in the chamber, and aerosol concentrations were only measured at five locations in addition to the measurements at the mouth of the recipient simulator. Aerosol concentration measurements also were only made at a height of 1.5 m. Thus, the average concentration data in Figure 6 should be interpreted with caution. Finally, the chamber used in our experiments was not ventilated in order to avoid the confounding factor of ventilation air currents during these initial studies. However, in real-world settings, ventilation is usually present, which produces air currents, encourages mixing and dispersion of aerosols, and removes aerosol particles from the room.

## CONCLUSIONS

Our study shows that universal masking consistently and significantly reduced the exposure of a recipient to respiratory aerosol particles produced by a source during coughing and breathing compared with experiments when the source and recipient were unmasked. These reductions were seen regardless of the orientation and separation distance between the source and recipient. When the source and recipient were unmasked, changes in orientation and separation distance affected the recipient exposure, but the effects of orientation and distance were reduced when both the source and recipient were masked.

## Data Availability

Experimental data is available upon request.

## ACKNOWLEDGEMENTS

We would like to thank the National Institute for Occupational Safety and Health (NIOSH) Morgantown maintenance, security, warehouse and housekeeping departments for their assistance and dedication during the ongoing pandemic. This work was supported by the US Centers for Disease Control and Prevention (CDC). The findings and conclusions in this report are those of the authors and do not necessarily represent the official position of NIOSH, CDC. Mention of any company or product does not constitute endorsement by NIOSH, CDC.

## Supplemental materials

**Figure S1:**
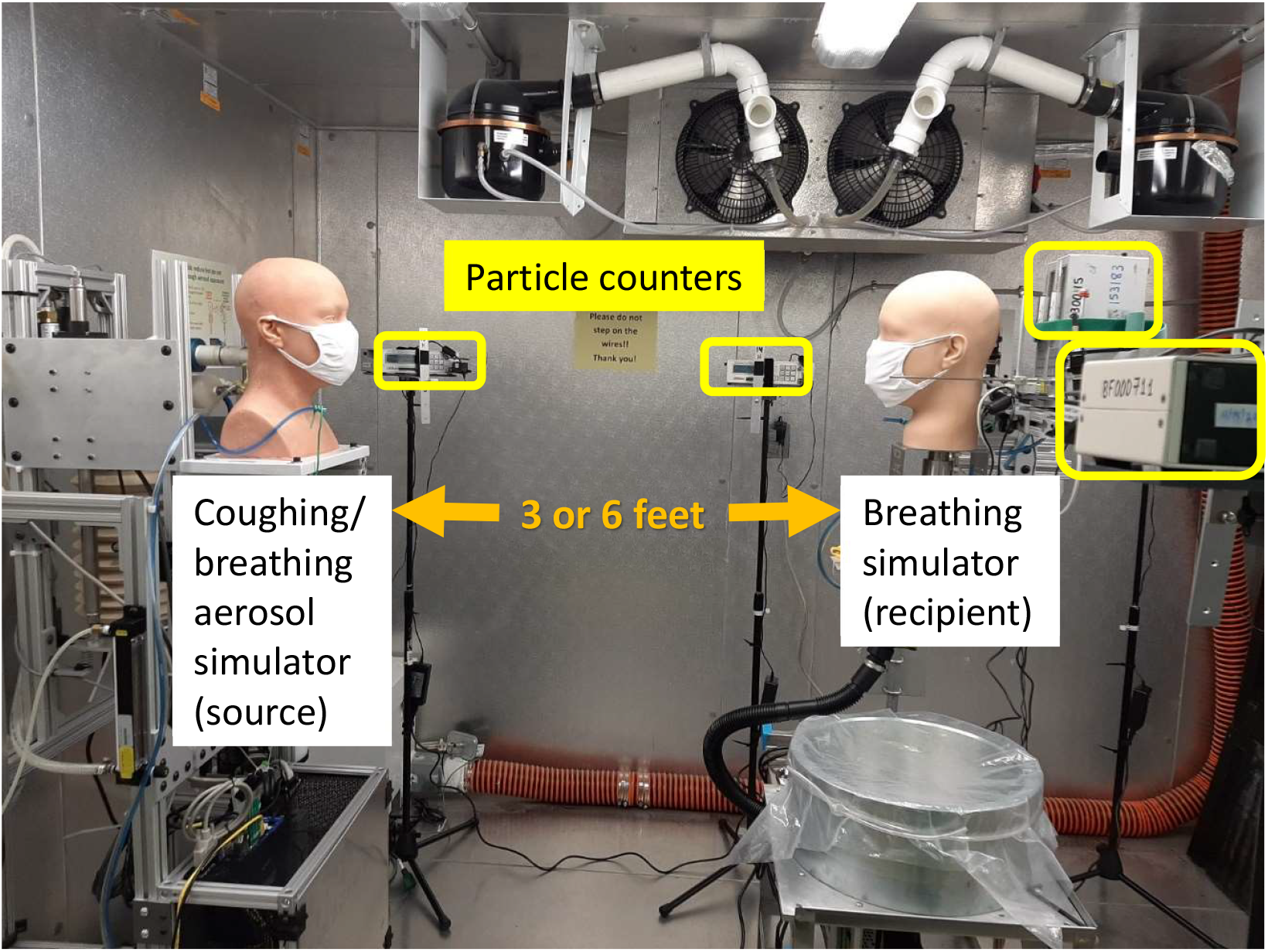
Photograph of experimental set-up in the environmental chamber. In this experiment, the source and recipient simulators were front-to-front 0.9 m apart.

**Figure S2:**
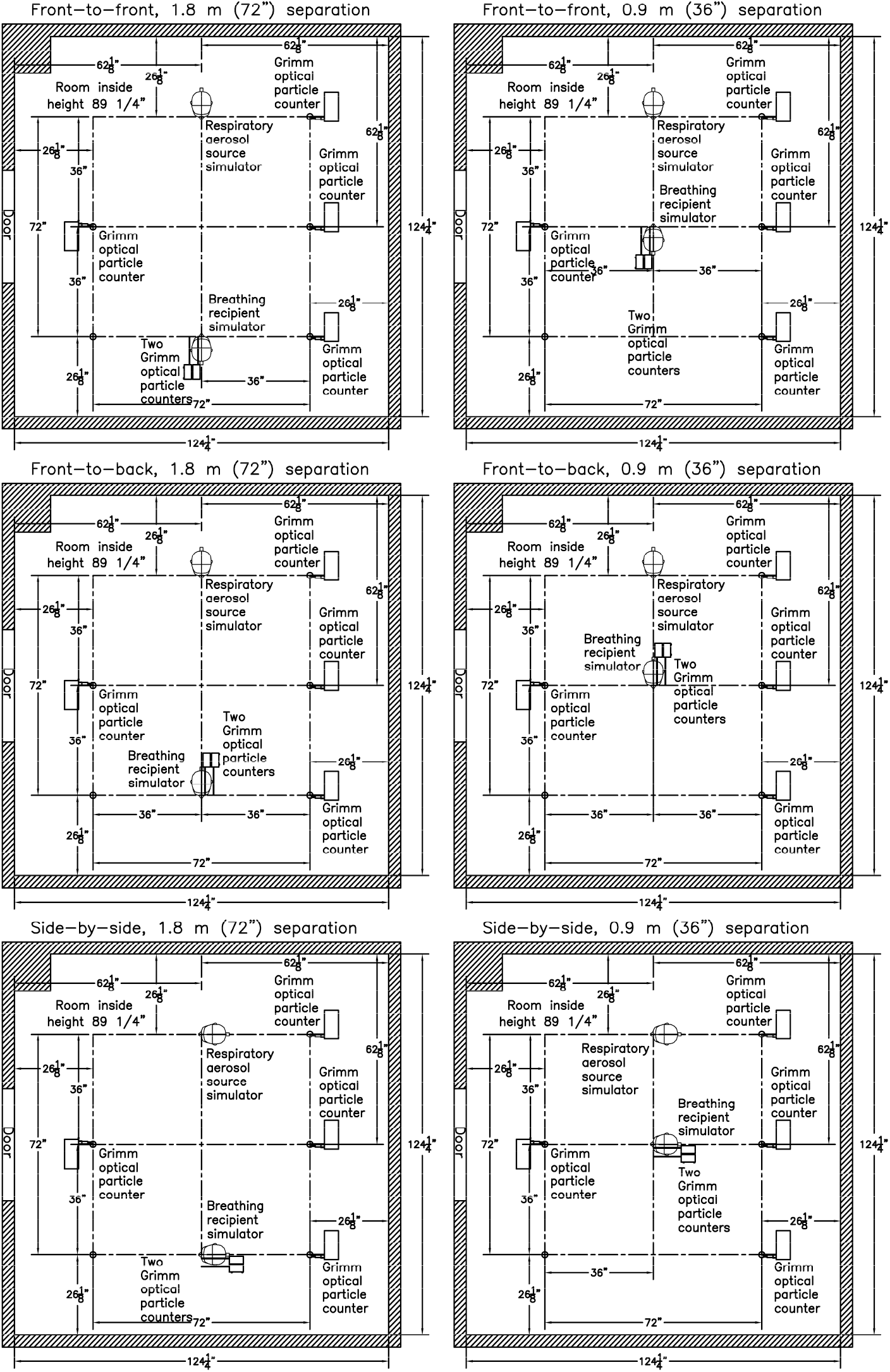
Layout of source simulator, recipient simulator and optical particle counters during experiments with different orientations and separation distances. Dimensions are in inches (1” = 2.54 cm). Drawings are to scale.

**Figure S3:**
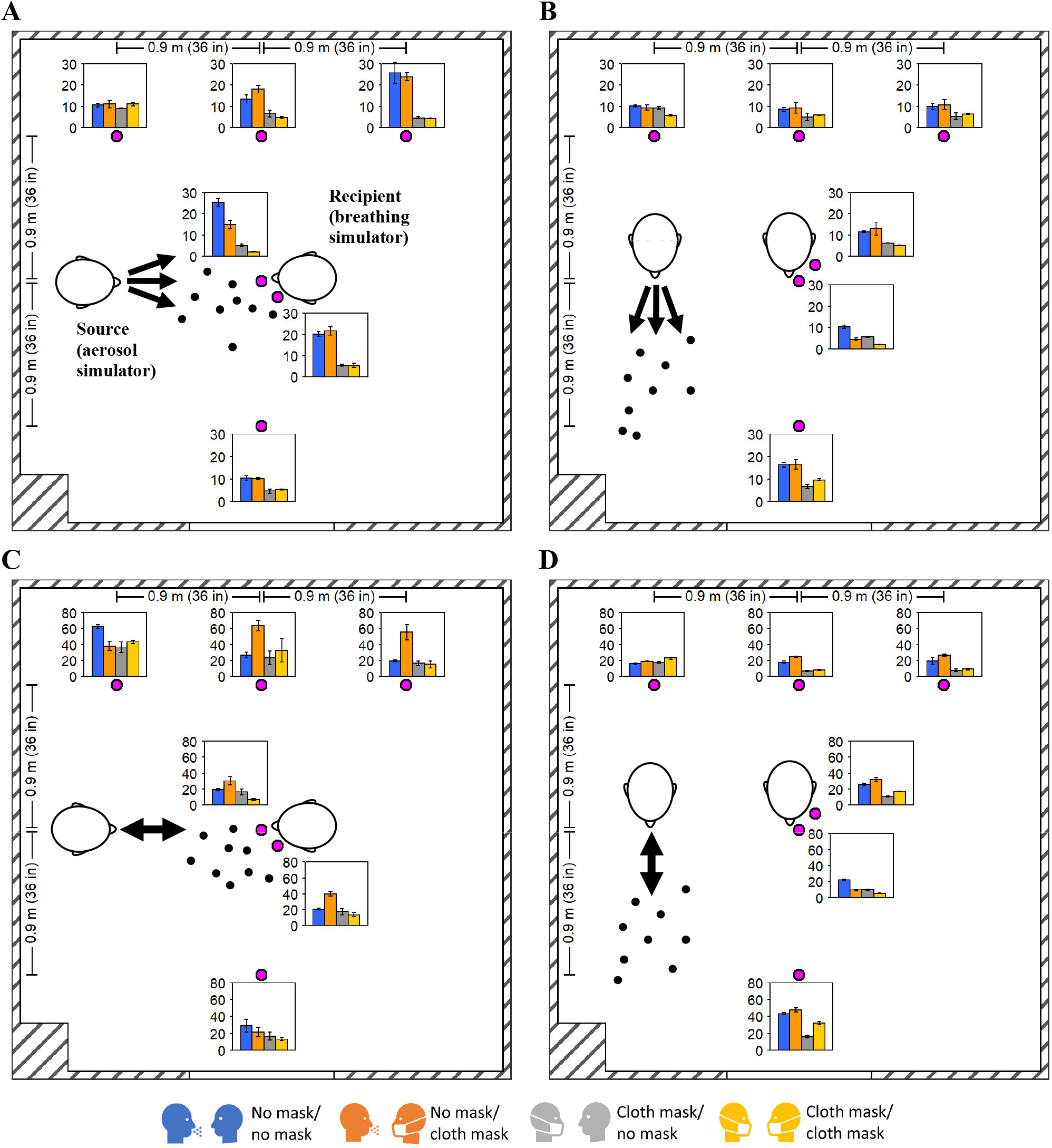
Mean aerosol concentrations at different locations in chamber with simulators 0.9 m apart. The magenta dots indicate the particle counter locations. The bar plots show the mean aerosol concentrations at each location with the source and recipient unmasked or masked as indicated in the legend below the plot. The mean values are reported in µg/m^3^ and were calculated over 15 minutes. Error bars indicate the standard deviations for three experiments. (A) Source is coughing, simulators are front-to-front. (B) Source is coughing, simulators are side-by-side. (C) Source is breathing, simulators are front-to-front. (D) Source is breathing, simulators are side-by-side.

**Figure S4:**
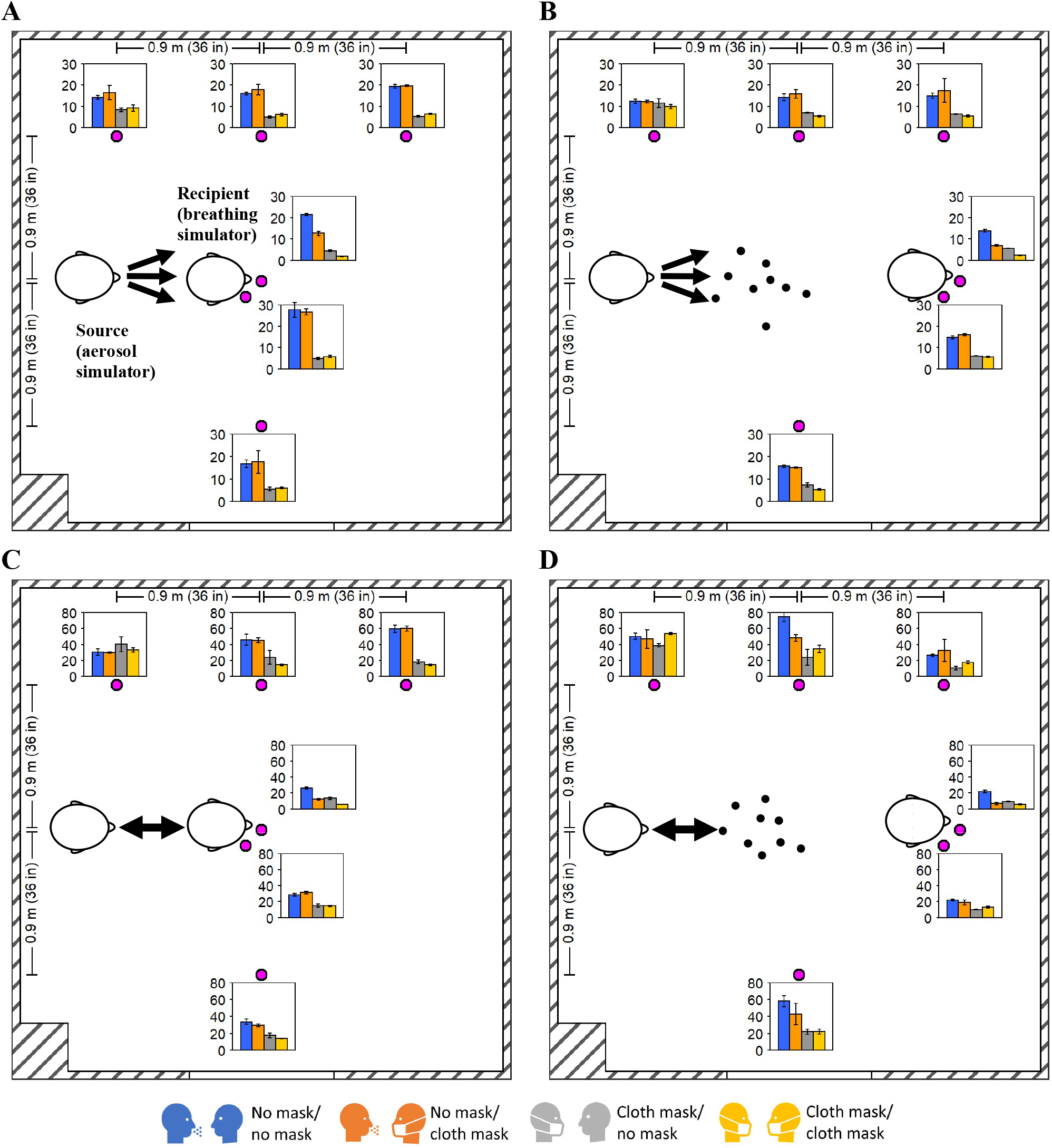
Mean aerosol concentrations at different locations in chamber with simulators front-to-back. The magenta dots indicate the particle counter locations. The bar plots show the mean aerosol concentrations at each location with the source and recipient unmasked or masked as indicated in the legend below the plot. The mean values are reported in µg/m^3^ and were calculated over 15 minutes. Error bars indicate the standard deviations for three experiments. (A) Source is coughing, simulators are 0.9 m apart. (B) Source is coughing, simulators are 1.8 m apart. (C) Source is breathing, simulators are 0.9 m apart. (D) Source is breathing, simulators are 1.8 m apart.

## REFERENCES

1. CDC: “How COVID-19 Spreads.” [Online] Available at https://www.cdc.gov/coronavirus/2019-ncov/prepare/transmission.html (accessed October 30, 2020).

2. Hamner, L., P. Dubbel, I. Capron et al.: High SARS-CoV-2 Attack Rate Following Exposure at a Choir Practice - Skagit County, Washington, March 2020. MMWR Morb. Mortal. Wkly. Rep. 69(19): 606–610 (2020).

3. Anderson, E.L., P. Turnham, J.R. Griffin, and C.C. Clarke: Consideration of the Aerosol Transmission for COVID-19 and Public Health. Risk Anal. 40(5): 902–907 (2020).

4. Morawska, L., and D.K. Milton: It Is Time to Address Airborne Transmission of Coronavirus Disease 2019 (COVID-19). Clin. Infect. Dis. 71(9): 2311–2313 (2020).

5. Ma, J., X. Qi, H. Chen et al.: COVID-19 patients in earlier stages exhaled millions of SARS-CoV-2 per hour. Clin. Infect. Dis. (online ahead of print)(2020).

6. Lavezzo, E., E. Franchin, C. Ciavarella et al.: Suppression of a SARS-CoV-2 outbreak in the Italian municipality of Vo’. Nature 584(7821): 425–429 (2020).

7. Moghadas, S.M., M.C. Fitzpatrick, P. Sah et al.: The implications of silent transmission for the control of COVID-19 outbreaks. Proc. Natl. Acad. Sci. U.S.A. 117(30): 17513–17515 (2020).

8. CDC: “Scientific Brief: Community Use of Cloth Masks to Control the Spread of SARS-CoV-2.” [Online] Available at https://www.cdc.gov/coronavirus/2019-ncov/more/masking-science-sars-cov2.html (accessed November 23, 2020).

9. Honein, M.A., A. Christie, D.A. Rose et al.: Summary of Guidance for Public Health Strategies to Address High Levels of Community Transmission of SARS-CoV-2 and Related Deaths, December 2020. MMWR Morb. Mortal. Wkly. Rep. 69(49): 1860–1867 (2020).

10. Howard, J., A. Huang, Z. Li et al.: An evidence review of face masks against COVID-19. Proc. Natl. Acad. Sci. U.S.A. 118(4): e2014564118 (2021).

11. Asadi, S., C.D. Cappa, S. Barreda, A.S. Wexler, N.M. Bouvier, and W.D. Ristenpart: Efficacy of masks and face coverings in controlling outward aerosol particle emission from expiratory activities. Sci. Rep. 10(1): 15665 (2020).

12. Davies, A., K.A. Thompson, K. Giri, G. Kafatos, J. Walker, and A. Bennett: Testing the efficacy of homemade masks: would they protect in an influenza pandemic? Disaster Med. Public Health Prep. 7(4): 413–418 (2013).

13. Milton, D.K., M.P. Fabian, B.J. Cowling, M.L. Grantham, and J.J. McDevitt: Influenza virus aerosols in human exhaled breath: particle size, culturability, and effect of surgical masks. PLoS Pathog. 9(3): e1003205 (2013).

14. Leung, N.H.L., D.K.W. Chu, E.Y.C. Shiu et al.: Respiratory virus shedding in exhaled breath and efficacy of face masks. Nat. Med. 26(5): 676–680 (2020).

15. Lawrence, R.B., M.G. Duling, C.A. Calvert, and C.C. Coffey: Comparison of performance of three different types of respiratory protection devices. J. Occup. Environ. Hyg. 3(9): 465–474 (2006).

16. Oberg, T., and L.M. Brosseau: Surgical mask filter and fit performance. Am. J. Infect. Control 36(4): 276–282 (2008).

17. Pan, J., C. Harb, W. Leng, and L.C. Marr: Inward and outward effectiveness of cloth masks, a surgical mask, and a face shield. Aerosol Sci. Technol. (online ahead of print)(2021).

18. Rothamer, D.A., S. Sanders, D. Reindl, and T.H. Bertram: Strategies to minimize SARS-CoV-2 transmission in classroom settings: Combined impacts of ventilation and mask effective filtration efficiency. MedRxiv (preprint) doi: 10.1101/2020.12.31.20249101(2021).

19. Lindsley, W.G., F.M. Blachere, B.F. Law, D.H. Beezhold, and J.D. Noti: Efficacy of face masks, neck gaiters and face shields for reducing the expulsion of simulated cough-generated aerosols. Aerosol Sci. Technol. 55(4): 449–457 (2021).

20. Lindsley, W.G., F.M. Blachere, D.H. Beezhold et al.: A comparison of performance metrics for cloth face masks as source control devices for simulated cough and exhalation aerosols. MedRxiv (preprint) doi: 10.1101/2021.02.16.21251850 (2021).

21. Brooks, J.T., D.H. Beezhold, J.D. Noti et al.: Maximizing Fit for Cloth and Medical Procedure Masks to Improve Performance and Reduce SARS-CoV-2 Transmission and Exposure, 2021. MMWR Morb. Mortal. Wkly. Rep. 70(7): 254–257 (2021).

22. Van Dyke, M.E., T.M. Rogers, E. Pevzner et al.: Trends in County-Level COVID-19 Incidence in Counties With and Without a Mask Mandate - Kansas, June 1-August 23, 2020. MMWR Morb. Mortal. Wkly. Rep. 69(47): 1777–1781 (2020).

23. Mitze, T., R. Kosfeld, J. Rode, and K. Walde: Face masks considerably reduce COVID-19 cases in Germany. Proc. Natl. Acad. Sci. U.S.A. 117(51): 32293–32301 (2020).

24. Wang, X., E.G. Ferro, G. Zhou, D. Hashimoto, and D.L. Bhatt: Association Between Universal Masking in a Health Care System and SARS-CoV-2 Positivity Among Health Care Workers. JAMA (2020).

25. Lyu, W., and G.L. Wehby: Community Use Of Face Masks And COVID-19: Evidence From A Natural Experiment Of State Mandates In The US. Health Aff. (Millwood) 39(8): 1419–1425 (2020).

26. Joo, H., G.F. Miller, G. Sunshine et al.: Decline in COVID-19 Hospitalization Growth Rates Associated with Statewide Mask Mandates - 10 States, March-October 2020. MMWR Morb. Mortal. Wkly. Rep. 70(6): 212–216 (2021).

27. Bergman, M.S., Z. Zhuang, D. Hanson et al.: Development of an advanced respirator fit-test headform. J. Occup. Environ. Hyg. 11(2): 117–125 (2014).

28. Lindsley, W.G., J.S. Reynolds, J.V. Szalajda, J.D. Noti, and D.H. Beezhold: A Cough Aerosol Simulator for the Study of Disease Transmission by Human Cough-Generated Aerosols. Aerosol Sci. Technol. 47(8): 937–944 (2013).

29. International Organization for Standardization: Respiratory protective devices -- Human factors -- Part 1: Metabolic rates and respiratory flow rates. (ISO/TS 16976-1:2015). [Standard] Geneva, Switzerland: ISO, 2015.

30. TSI: Penetration of N95 Filtering Facepiece Respirators by Charged and Charge-neutralized Nanoparticles, by Han, H.-S., and M. Prell. Report No. Application Note RFT-007 (US). Shoreview, MN USA: TSI 2010.

31. TSI: Fit testing using size-selected aerosol, by Halvorsen, T. Report No. Application Note ITI-062. Shoreview, MN USA: TSI 1998.

32. TSI: PortaCount Pro 8030 and PortaCount Pro+ 8038 Respirator Fit Testers Operation and Service Manual, by TSI. Report No. P/N 6001868, Revision p. Shoreview, MN USA: TSI 2015.

33. Janssen, L., and R. McKay: Respirator performance terminology. J. Occup. Environ. Hyg. 14(12): D181–D183 (2017).

34. R Core Team: “R: A language and environment for statistical computing.” [Online] Available at https://www.R-project.org/ (accessed March 22, 2021).

35. Lenth, R.V.: Least-Squares Means: The R Package lsmeans. Journal of Statistical Software 69(1): 33 (2016).

36. Kuznetsova, A., P.B. Brockhoff, and R.H.B. Christensen: lmerTest Package: Tests in Linear Mixed Effects Models. Journal of Statistical Software 82(13): 26 (2017).

37. Verma, S., M. Dhanak, and J. Frankenfield: Visualizing the effectiveness of face masks in obstructing respiratory jets. Phys. Fluids (1994) 32(6): 061708 (2020).

38. Fennelly, K.P.: Particle sizes of infectious aerosols: implications for infection control. Lancet Respir. Med. 8(9): 914–924 (2020).

39. Gralton, J., E. Tovey, M.L. McLaws, and W.D. Rawlinson: The role of particle size in aerosolised pathogen transmission: a review. J. Infect. 62(1): 1–13 (2011).

40. Lindsley, W.G., J.D. Noti, F.M. Blachere, J.V. Szalajda, and D.H. Beezhold: Efficacy of face shields against cough aerosol droplets from a cough simulator. J. Occup. Environ. Hyg. 11(8): 509–518 (2014).

41. Bourouiba, L., E. Dehandschoewercker, and John W.M. Bush: Violent expiratory events: on coughing and sneezing. J. Fluid Mech. 745: 537–563 (2014).

42. Bahl, P., C. Doolan, C. de Silva, A.A. Chughtai, L. Bourouiba, and C.R. MacIntyre: Airborne or droplet precautions for health workers treating COVID-19? J. Infect. Dis. (online ahead of print)(2020).

